# Self-Stigma And Vicarious Stigma Experienced By Parents Of Children With Neurodevelopmental Disorders Living In Rivers State, Nigeria

**DOI:** 10.1101/2024.10.31.24316480

**Authors:** Onunze Chukwunonso, Ohakpougwu Chukwuebuka, Steven Isioma, Obilor Sandra, Emezuo Theresa

## Abstract

**Background:** parents of children living with Neurodevelopmental Disorders (NDDs) continue to be shamed despite the herculean task of caring for these children. Under the weight of this “double burden” parents experience double stigma: internalized and public with consequent mental health challenges.

**Aim:** This study was conducted to explore the prevalence of self and vicarious stigma among parents of children with NDDs in Rivers State, Nigeria, as well as the correlations of these stigma with depression among these parents.

**Methods:** This was a cross-sectional study of 62 parents in Port Harcourt, Nigeria employing quantitative data collection methods. Data was collected via semi-structured interviewer administered questionnaires and analysed using SPSS version. Data was summarized using proportions, mean, standard deviations. Chi square and Spearman’s correlation test were done and the level of significance was pre-determined at 5% (p<0.005).

**Results:** Autism Spectrum Disorder (ASD) was the commonest primary diagnosis of children in the study centres at 62.9 %. The mean age of parents was 36.4+/-7.6 years. About half of parents experienced self-stigma (50.0 %), whereas 61.3 % of parents experienced sadness due to vicarious stigma and 54.8 % experienced anger due to vicarious stigma. Overall, 56.5 % of parents suffered from depression.

**Conclusion:** Vicarious stigma which is public stigma on parents of children with NDDs overshadowed self-stigma. This makes urgent the need for more public education on ways society can destigmatize NDDs and provide more support for parents. Across board, stigma in any form is present can cause depression in parents, hence there is need for long term psychosocial support for these parents.

## INTRODUCTION AND BACKGROUND

Neurodevelopmental disorders are disabilities associated primarily with the functioning of the neurological system and brain [1]. They include autism spectrum disorders (ASD), motor disorders, tic disorders communication disorders and specific learning disorders (SLD) or attention deficit/ hyperactivity disorder (ADHD)[2]. Although once thought of as disorders of childhood, it is becoming increasingly apparent that the majority of these disorders persist over the course of one’s lifetime. Parents caring for children with NDDs experience more medical, psychological, and familial dysfunction because of the additive nature of children’s NDDs and disease manifestations, leading to long-term exposure to stress and caring responsibilities [3]. Parents experience higher levels of stress with NDDs, particularly from children’s emotional and behavioural challenges than with other developmental disabilities [4]. Children with neurodevelopmental disorders require frequent supports from inpatient and outpatient medical, and community-based systems, including primary care, emergency care, rehabilitation, and government disability support [5].

Knowledge and awareness about ASD have important implications for help-seeking behaviour. In cases where etiology of ASD is attributed to spiritual causes as reflected by study findings, [6] it is obvious that the first point of contact in the process of help-seeking would be traditional or religious healers, a practice commonly observed among children with ASD in Nigeria. Stigma and derogatory comments from neighbours have actually made many parents with children with ASD to lock up or hide such children away in their homes, thus depriving them of needed early interventions that have been shown to improve prognoses in many cases. [7]

The paucity of precise epidemiological data on NDD in low-and-middle income countries has impeded the development of public health interventions and also adequate support for parents/caregivers who bear the burden of nurturing affected children.

Stigma is central to the experience of parenting a child with a neurodevelopmental disorder. Stigma is a multifaceted construct and is commonly described in terms of stereotypes, prejudice and discrimination [8].

Stigma ascribed to children with neurodevelopmental disorders is often transferred to their parents. Societal prejudice and discrimination, reinforced by stereotypes which manifests as public stigma and loss of self-esteem which manifests as self-stigma can negatively affect the mental health of parents raising a child with neurodevelopmental disorder. Self-stigma is an internalized form of bias that is often experienced by parents of children with neurodevelopmental disorders. This type of stigma can manifest in various ways, such as feeling shame, guilt, and embarrassment, or a general sense that they are inadequate parents [9]. This can be further complicated by parent’s negative feelings towards the diagnosis, and the fear of not being able to do enough to help their children. As a result, many parents may retreat emotionally, or avoid seeking treatment or support to help them deal with their own needs. When these stereotypes are internalized, there is increased depression, diminished quality of life. Beyond self-stigma, parents of children with neurodevelopmental disorders also experience vicarious stigma, which is borne out of their relationship with the child. Parents may experience ‘vicarious stigma’, a tendency to worry about the impact of stigma on their child as they help their children with NDD navigate the interpersonal and social difficulties [10]. This manifests as feelings of guilt, sadness, anger, or frustration when their child is excluded from birthday parties or the school team for instance.

Parents of children with NDD can experience depressive symptoms as a result of stigma. The effect of Vicarious and self-stigma experienced by parent of children with NDD and discovered that Parents with greater Self Stigma were found to show more depression and lower Quality of Life (QoL)[11].

In the face of stigma associated with parenting children with NDDs parents can often adopt various methods of coping such as secrecy coping. Secrecy coping is a phenomenon that is often observed and experienced by parents of children with neurodevelopmental disorders. While secrecy may be seen as a coping mechanism to protect the feelings of the child, the parent or both, it can lead to a range of issues, including isolation, lowered self-esteem, and difficulty in obtaining support from family and friends [12].

Studies have shown that secrecy, while sometimes necessary, can be isolating and can interfere with the ability of parents to access and receive support, as well as to process their emotions [13]. Financial concerns can also be a factor in the secrecy that a parent may experience. Many of the treatments and therapies available for these children can come at a great cost, and these financial needs can lead to a sense of isolation and secrecy, as the parent worries about how the family will afford the necessary services for their child [14].

Given the gravity of the challenge of taking care of children with NDD, and the consequent stigma, it is now a public health emergency to survey affected parents and develop interventions tailored to their needs. Hence, this study is intended to investigate in-depth the prevalence of both self and vicarious stigma among parents of children with NDD in a low resource setting, and proffer sustainable interventions and advocacy.

The findings from the study will contribute to the understanding of Neurodevelopmental disorders within the Nigerian cultural setting and low-and-middle-income countries by extension. Furthermore, this will support ongoing efforts to understand and diminish the stigma of Neurodevelopmental disorders and possibly improve the level of acceptance, sensitivity, treatment, and support for individuals affected by Neurodevelopmental disorders.

## MATERIALS AND METHODS

### Study setting

The research setting was the parents of children with NDD (Neurodevelopmental disorders) who visit Blazing heart Autism centre and associated centres where children with NDD are being managed within Port Harcourt and Obio/Akpo LGA in Rivers state.

### Site of study

Interested participants were contacted through Blazing Heart Autism Centre and other advocacy groups and centres across Port Harcourt through phone calls, email or in actual BHAC office.

Also, another study site were University of Port Harcourt Teaching Hospital (UPTH) Neurology and Mental Health unit.

### Study Population

Participants in this study were at least 18 years of age and a custodial parent of a child with mental health and/or neurodevelopmental disorders between the ages of 2 and 14 years. For the purposes of this study, the child’s mental health and/or neurodevelopmental disorder included ADHD, anxiety disorders, autism spectrum disorder, bipolar and related disorders, conduct disorder, depressive disorders, oppositional-defiant disorder, and post-traumatic stress disorder.

### Study Size

#### Sample Size

For the purposes of this study, a sample size of 62 participants who are parents of at least a child with Neurodevelopmental disorder were used.Participants are recruited using criterion sampling. Participants with certain characters defined by the exclusion and inclusion criteria were used for the study

#### Inclusion Criteria

1. Parents and caregivers of a child with NDD for at least 50% of the time
2. Child with NDD must be 2 to 14 years of age
3. The parent or caregiver and the child must be living within Port Harcourt and Obio/Akpor LGA

#### Exclusion Criteria

1. Parents and caregivers of a child with NDD for less than 50% of the time
2. Parents and caregiver of Child with NDD who is less than 2 years and more than 14 years of age
3. The parent or caregiver and the child who lives outside of Rivers State
4. Parents of children with acquired Neurological disorders.

### Source of Data

Primary source of data was obtained from a semi-structured questionnaire completed by respondents. The data were transferred to an electronic spreadsheet and stored for analysis.

### Survey Style

Research participants completed measures of demographics, vicarious stigma, Self stigma, secrecy coping, and depression. Internal consistency for each measure were calculated for the sample in this study and reported. Demographic factors were selected that correspond with existing stigma research literature and reflect parent descriptors and will include year of birth, gender, ethnicity, race, marital status, employment status, yearly family income, highest level of achieved education, and custodial relationship to their child (e.g., biological parent, step-parent, adoptive parent). We also collected demographics that were specific to questions of parenting children with mental health and/or neurodevelopmental disorders. These included items about the child included year of birth, gender, primary diagnoses, approximate year of diagnosis, professional who provided the diagnosis, and type of treatments/supports the child was currently receiving. An already developed Vicarious Stigma Scale (VSS) were used for this study [11]. We constructed a vignette-based items representing hypothesized experience that yielded vicarious stigma to which respondents provided responses on Likert scales. The VSS were used for the study. Participants were instructed to respond to each situation with two 10-point scales: “How much does this make you feel sad…?” or “How much does this make you feel angry…?” (10 = very much). Scores were summed to yield individual vicarious stigma sad and anger scores. Remaining measures will then be selected to represent key constructs to which vicarious stigma were compared: SS, secrecy, and depression. Self-stigma was assessed using the Parents’ Self-Stigma Scale. This 11-item, self-report measure asks participants to rate items on five-point agreement scales (5 = almost all the time). Total score were computed by calculating the sum of items. The PSSS has demonstrated sufficient reliability for this Secrecy coping were measured using an adapted and expanded version of the 5-item Secrecy Coping Scale [12]. This measure is adapted by rewording phrases about an adult’s mental illness to reflect the experiences of parents of children with mental health and/or neurodevelopmental disorders (e.g., “For me to be successful at work, it is best to hide my diagnosis or problems.” was changed to “For my child to be successful at school, it is best to hide his/her diagnosis or problems.”). Participants were instructed to rate each item on a 6-point Likert scale (6 = strongly disagree). Half the items were reverse scored. Total scores were computed by calculating the sum of items. Lower scores represent greater levels of secrecy coping, and less promotion of disclosure. Depression was measured using the Centre for Epidemiological Studies Short Depression Scale (CESD-R-10). This 10-item, self-report measure asks participants to respond to items (e.g., “I felt depressed”) using a rating scale (rarely or none of the times (less than 1 day) to all of the time (5–7 days). The total score was computed by calculating the sum of items. Higher scores represent greater levels of depression.

## RESULT

### Socio-demographic characteristics

a total of 62 parents participated in the study. The mean age of the respondents was 36.4±7.6 years. Majority, (40.3%) were in the 30 to 39 years age group. Most of the parents had completed a tertiary education (82.3%) at the time of this study, are currently married (87.1%), employed (95.2%), and earn less than 50,000 naira monthly (33.9%). Fifty-eight percent of the parents had at least a child under 5 years (58.1%), diagnosed of autism spectrum disorder (ASD) (62.9%), and on individual child therapy (79.0%). All the respondents were the biological parent of the child and have been the primary care giver of the child for at least 50% of the child’s life (Table 1).

### Sadness due to vicarious stigma

To assess the level of sadness due to vicarious stigma, seven Likert scale questions were asked with a zero (0) to 10 possible responses. Zero (0) meaning “no sadness due to vicarious stigma” and 10 meaning “very much sadness due to vicarious stigma”. The mean score of the responses was used to dichotomize sadness level into low and high. Respondents whose scored below the mean were regarded as experiencing low level of sadness due to vicarious stigma while those who scored the mean number and above were classified as experiencing higher sadness.

The mean score of sadness caused by vicarious stigma is 24.2±16.7. Thirty-eight respondents scored 24 and above thus were regarded to having high sadness due to vicarious stigma (Table 2).

### Anger due to vicarious stigma

To ascertain the level of anger caused by vicarious stigma, seven Likert scale questions were asked with a zero (0) to 10 possible responses. Zero (0) meaning “vicarious stigma does not make me Angry” and 10 means vicarious stigma causes “very much Anger”. The mean score of the responses was used to classify the level of anger into low and high levels. Respondents who scored below the mean were regarded as experiencing low level of anger due to vicarious stigma while those who scored the mean value and above were classified highly angered from vicarious stigma.

The mean score for the level of anger caused by vicarious stigma is 19.0±15.8. The majority (54.8%) of respondents scored the mean value and above thus were referred to as experiencing very much anger due to vicarious stigma (Table 2).

### Self-stigma

Self-stigma was measured using an 11 item Likert scale questions. The possible responses were one to five. The mean value of the responses was obtained and used to classify the respondents into experiencing lesser or greater self-stigma.

The mean score for self-stigma is 26.7±6.5. Thirty-one (50.0%) of respondents experience both lesser and greater levels of self-stigma (Table 2).

### Secrecy Coping

Secrecy coping was assessed using six Likert scale questions. The response was pegged at one to six. The mean score of the responded where used to dichotomize the secrecy coping into higher and lower level. Parents who scored below the mean were regarded as having lower secrecy coping while those who scored the mean number and above classified as having higher secrecy coping.

The mean score for the level of secrecy coping is 15.8±6.0. Only 48.4% of the parents had a lower secrecy coping as shown in Table 2.

### Depression

To measure parents’ level of depression, 10 Likert scale questions were asked with one to four possible responses. One represents lower depression while four represents greater depression levels. The mean score of the responses was used to classify depression as in above. Respondents whose score below the mean was regarded as having lower level of depression while those who scored the mean number and above classified as experiencing higher depression.

The mean score for the level of secrecy coping is 24.1±6.3. Fifty-seven percent of the parents **experience** a high level of depression as shown in Table 2 below

## ASSOCIATIONS

### Sadness due to vicarious stigma

Thirty-eight (61.3%) respondents experience high levels of sadness due to vicarious stigma. Secrecy coping and child primary diagnosis were significantly associated with sadness due to vicarious stigma. Respondents with higher secrecy coping are three times more likely to experience sadness due to vicarious stigma than parents with lower secrecy coping (COR = 3.43, CI= 1.17 – 10.04) table 3.

### Anger due to vicarious stigma

Out of the 62 respondents, thirty-four (54.8%) respondents experience high levels of anger due to vicarious stigma. For a test of significance to identify factors associated with anger due to vicarious stigma, the Chi-square test was used at *P*=0.05 (Table 4). Depression, respondents age and employment status were found to be significantly associated with anger due to vicarious stigma. Parents with greater depression are four times more likely to experience anger due to vicarious stigma (COR=3.709, CI = 1.29 – 10.69) than their counterpart with lower level of depression. Age of the respondents was also found to be significantly associated with anger due to VS, parents who were 40 years and above are six times more likely to experience anger due to VS than parents less than 30 years of age (COR = 5.893, CI= 1.14 – 25.23).

### Self-stigma

Fifty percent of the respondents reported that they experience a high level of self-stigma. Table 5 shows the result from chi-square test to identify factors associated with self-stigma. Only family income level was found to be significant. Parents who earn between one hundred and one to five hundred-thousand-naira per month are 80% times less likely to experience self-stigma than parents who earn fifty thousand naira and below monthly (COR = 0.164, CI= 0.04 – 0.67).

## DISCUSSION

Globally, self and vicarious stigma continue to plague the mental health of parents and caregivers of children living with NDD. Our study re-emphasised the huge negative impact of self and vicarious stigma on the mental health of parents of children living with NDD. More than half of respondents in our study reported experiencing vicarious stigma which corresponds with a study in Lagos but contrasts sharply with a study in U.S study which reported about 20 percent of respondents experiencing vicarious stigma [9]. Findings from our study reveal that half of respondents experienced self-stigma, which was equivalent to studies in China and Virginia [15]. This differed markedly with the study in Lagos which showed that 80 percent of parents self-stigmatize [9]. This finding agrees with findings in Hong Kong, where self-stigma was sky-high among respondents. In our study, parents who reported feeling greater levels of self-stigma had higher levels of depression as opposed to those who reported lower levels of self-stigma. This finding is corroborated by other studies exploring self-stigma in parents of children with NDD [12]. However, our research showed that parents with anger due to Vicarious stigma had higher levels of depression as opposed to parents experiencing sadness due to vicarious stigma, whilst parents experiencing self-stigma had lower levels of depression as compared to the previous two groups. This appeared to differ from the findings in Chicago which reported much higher levels of depression among parents experiencing sadness due to vicarious stigma than all other categories of stigma [11]. Even though secrecy coping might help the child affected from being stigmatized by the public, it can have deleterious effects on parent’s mental health. In this study about half of respondents admitted to coping via secrecy. Following this, about half of respondents had high levels of self-stigma, and the same number had high level of depression. It is no coincidence that same percentage of parents who subscribed to secrecy coping also experience worse mental health. This agreed with findings of a Chicago study where parents who were inclined to cope via secrecy reported higher depression whilst those more likely to disclose had better quality of life [11]. Our study made significant findings on the influence of family’s income on self-stigma. Here, families who earned the least /lower incomes experienced far greater self-stigma as opposed to those who were high income earners. This disparity was far significant statistically as well as opposed to a mixed methods study in Lagos where family income had no significant influence on self-stigma regardless of family’s income [9]. Despite global knowledge of the effect of stigma on the mental health of parents of children living with NDDs, there has been little progress in advancing methods and strategies to deal with internalized and vicarious stigma parents feel. Generally, psychoeducation, cognitive therapy, as well as peer support have been proposed and practiced in the western world with varying degrees of success. Nevertheless, in developing countries there is a lacuna with regards to interventions. One simple intervention could be creating more avenues to combat secrecy coping as well as increasing public education on stigma parents of children with NDD face, and the publics role in compounding this problem.

### Strengths and limitations of study

This study contributes to the pertinent discussion on stigma faced by parents with children with NDDs especially in developing countries where there is a paucity of studies on this subject. Employing mixed method study design in core centres for children with NDDs, ensured that respondents were directly impacted by caring for such children, as well as generally increased rigorous data collection and upholds scientific quality. Nevertheless, since this is a cross-sectional study, the generalizability of findings may be limited, and causality cannot be readily inferred. Also, the one directional phrasing of our Likert statements has a likelihood of increasing response-set bias. Considering small sample size, and sampling method there could also have been some “clustering effect”. More large studies are needed especially in developing countries to further study the effect of stigma on the mental health of parents of children with NDD to push policy in the right direction and concretely related self-stigma to poor quality of life for such parents. Importantly, there is a need to find definite solutions for parents experiencing self and vicarious stigma.

## Conclusion

The stigma surrounding NDDs continue to make them a taboo topic and a cause of shame, sadness, anger and depression among parents of children suffering from this health challenge. Not enough is being done to raise awareness of the ills that stigma perpetuates on the mental health of parents and the time is right to change the narrative. Hospitals and health centres offering services to children with NDDs should also offer counselling, psychotherapy, and peer support activities to parents of such children. Social welfare units at such facilities can also offer financial aid to parents who earn a low income to help alleviate some of the burden for care. In counselling and psychotherapy sessions, it is important to help reorientate parents to address internalized or enacted stigma head-on and teach good coping strategies.

Caring for children with NDDs is a herculean task, thus keeping an open-door policy, and a log of their emotions that they can discuss in therapy sessions. Research exploring the efficacy of various approaches to curtail self and vicarious stigma can help contribute to an arsenal of evidence-based methods to ease the long suffering of these parents. This will be a great relieve to thousands globally and will guarantee quality for children with NDDs worldwide.

## Data Availability

All data produced in the present study are available upon reasonable request to the authors

## References

1. Cainelli E, Bisiacchi P. Neurodevelopmental Disorders: Past, Present, and Future. Children (Basel). 2022 Dec 24;10(1):31. doi: 10.3390/children10010031. PMID: 36670582; PMCID: PMC9856894.

2. Deb, S.S., Roy, M., Bachmann, C., et al. Specific Learning Disorders, Motor Disorders, and Communication Disorders. Textbook of Psychiatry for Intellectual Disability and Autism Spectrum Disorder. Bertelli, M.O., Deb, S. et al (eds). 2022 Springer, Cham. 10.1007/978-3-319-95720-3_18

3. Lach, L. M., Kohen, D. E., Garner, R. E., Brehaut, J. C., Miller, A. R., Klassen, A. F., & Rosenbaum, P. L. (2009). The health and psychosocial functioning of caregivers of children with neurodevelopmental disorders. Disability and Rehabilitation, 31, 741– 752.

4. Craig F, Margari F, Legrottaglie AR, Palumbi R, de Giambattista C, Margari L. A review of executive function deficits in autism spectrum disorder and attention-deficit/hyperactivity disorder. Neuropsychiatr Dis Treat. 2016 May 12;12:1191–202. doi: 10.2147/NDT.S104620. PMID: 27274255; PMCID: PMC4869784.

5. Currie G, Szabo J. Social isolation and exclusion: the parents’ experience of caring for children with rare neurodevelopmental disorders. Int J Qual Stud Health Well-being. 2020 Dec;15(1):1725362. doi: 10.1080/17482631.2020.1725362. PMID: 32048917; PMCID: PMC7034477.

6. Bakare MO, Taiwo OG, Bello-Mojeed MA, Munir KM. Autism Spectrum Disorders in Nigeria: A Scoping Review of Literature and Opinion on Future Research and Social Policy Directions. J Health Care Poor Underserved. 2019;30(3):899–909. doi: 10.1353/hpu.2019.0063. PMID: 31422978; PMCID: PMC6815667

7. Weiss MG, Ramakrishna J. Stigma interventions and research for international health. Lancet. 2006 Feb 11;367(9509):536–8. doi: 10.1016/S0140-6736(06)68189-0. PMID: 16473134.

8. Corrigan P.W., Kerr A., Knudsen L. (2005). The stigma of mental illness: Explanatory models and methods for change. Applied and Preventive Psychology, 11(3), 179–190.

9. Oduyemi, Aminat & Okafor, Ifeoma & Eze, Ugochukwu & Akodu, Babatunde & Roberts, Alero. (2021). Internalization of stigma among parents of children with autism spectrum disorder in Nigeria: a mixed method study. BMC Psychology. 9. 10.1186/s40359-021-00687-3.

10. Corrigan, P. W., Watson, A. C., & Miller, F. E. (2006). Blame, shame, and contamination: The impact of mental illness and drug dependence stigma on family members. Journal of Family Psychology, 20(2), 239–246. 10.1037/0893-3200.20.2.239

11. Serchuk, M. D., Corrigan, P. W., Reed, S., & Ohan, J. L. (2021). Vicarious stigma and self-stigma experienced by parents of children with mental health and/or neurodevelopmental disorders. Community Mental Health Journal, 57(8), 1537– 1546. 10.1007/s10597-021-00774-0

12. Eaton, K., Ohan, J. L., Stritzke, G. K., & Corrigan, P. W. (2016). Failing to meet the good parent ideal: Self-stigma in parents of children with mental health disorders. Journal of Child and Family Studies, 25(10), 3109–3123. 10.1007/s10826-016-0459-9.

13. Smith I. C., Edelstein J. A., Cox B. E., White S. W. (2018). Parental disclosure of ASD diagnosis to the child: A systematic review. Evidence-Based Practice in Child and Adolescent Mental Health, 3(2), 98– 105. 10.1080/23794925.2018.1435319

14. Saunders BS, Tilford JM, Fussell JJ, Schulz EG, Casey PH, Kuo DZ. (2015) Financial and employment impact of intellectual disability on families of children with autism. Fam Syst Health.33(1):36–45. doi: 10.1037/fsh0000102. Epub 2015 Jan 12. PMID: 25581557; PMCID: PMC4355223.

15. Winnie W.S. Mak, Yvonne T.Y. Kwok. Internalization of stigma for parents of children with autism spectrum disorder in Hong Kong, Social Science & Medicine, Volume 70, Issue 12, 2010, Pages 2045–2051, ISSN 0277-9536. 10.1016/j.socscimed.2010.02.023.

